# Inferring the true number of SARS-CoV-2 infections in Japan

**DOI:** 10.1101/2022.04.01.22273214

**Authors:** Lauren McKenzie, Affan Shoukat, Kai On Wong, Koju Itahashi, Eiji Yasuda, Alex Demarsh, Kamran Khan

## Abstract

**Introduction:** In Japan, as of December 31, 2021, more than 1.73 million laboratory-confirmed cases have been reported. However, the actual number of infections is likely to be under-ascertained due to the epidemiological characteristics such as mild and subclinical infections and limited testing availability in the early days of the pandemic. In this study, we infer the true number of infections in Japan between January 16, 2020, and December 31, 2021, using a statistical modelling framework that combines data on reported cases and fatalities.

**Methods:** We used reported daily COVID-19 deaths stratified into 8 distinct age-groups and age-specific infection fatality ratios (IFR) to impute the true number of infections. Estimates of IFR were informed from published studies as well seroprevalence studies conducted in Japan. To account for the uncertainty in IFR estimates, we sampled values from relevant distributions.

**Results:** We estimated that as of December 31, 2021, 2.90 million (CrI: 1.77 to 4.27 million) people had been infected in Japan, which is 1.68 times higher than the 1.73 million reported cases. Our meta-analysis confirmed that these findings were consistent with the intermittent seroprevalence studies conducted in Japan.

**Conclusions:** We have estimated that a substantial number of COVID-19 infections in the country were unreported, particularly in adults. Our approach provides a more realistic assessment of the true underlying burden of COVID-19. The results of this study can be used as fundamental components to strengthen population health control and surveillance measures.

## Introduction

Reliable estimates of the true number of SARS-CoV-2 infections in a population are necessary for evaluating the course of COVID-19, effectiveness of control strategies, and situational awareness. The number of confirmed cases, while readily available, tends to largely underestimate the true number of infections due to several characteristics of SARS-CoV-2 including mild and subclinical infections [1,2], high transmissibility [3–5], and an incubation period with a long-tailed distribution [6]. Additionally, limited availability of tests, imperfect test sensitivities, and delays in reporting further exacerbates the under-ascertainment of infections.

In Japan, as of December 31, 2021, more than 1.73 million laboratory-confirmed cases have been reported since the first identified case [7]. However, studies based on seroprevalence data in Japan have reported a high degree of under-ascertainment of cases [8–12]. For instance, the ascertainment rate of non-severe cases was estimated to be 0.44 at the start of the pandemic [12]. A more recent study found that actual infection numbers were 10-fold higher than those reported by PCR testing in the city of Kobe [9]. Seroprevalence studies generally provide more accurate estimation of population prevalence. However, these studies are sparse in time and space and their use is not recommended in low prevalence settings as results could lead to a false sense of security regarding the extent of immunity in the population [13] impeding mitigation strategies and interventions. In addition, tests conducted before seroconversion or after antibodies have waned can lead to false positives and false negatives [14].

To estimate an accurate level of case ascertainment in Japan, we developed a statistical modelling framework that combines data on reported cases and deaths. We posit that reported death counts are a more accurate indicator of disease burden than confirmed cases [15,16]. Countries with highly accessible healthcare and reliable collection of COVID-19 mortality data, including Japan, routinely and comprehensively capture and report disease-specific deaths [15,17]. Accordingly, we used age-specific infection fatality ratios (IFRs) along with reported death counts to infer the true number of infections in the country. The IFR value represents the proportion of deaths among all individuals infected with COVID-19, including symptomatic and subclinical infections. Consequently, our methodology enables us to remove selection bias due to individuals with mild symptoms or asymptomatic infection in case ascertainment. Our results can inform the efficacy of public health measures taken to protect the population from infection and estimate the underlying population immunity with higher accuracy and confidence than measures based on reported cases.

## Methods

The total number of infections was estimated using daily reported COVID-19 deaths in Japan between January 16, 2020 and December 31, 2021 [7]. The number of deaths *D*_*t*_ on each day *t* was split into 8 distinct age groups using estimated proportions of COVID-19 deaths by age in Japan [18]. Estimates of IFR were informed from published seroprevalence studies (Table 1). For each age-group, the number of infections *N*_*t*_ associated number of deaths *D*_*t*_ on day *t* was imputed using the formula

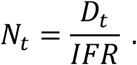

**Table 1.** Proportion of COVID-19 deaths in Japan, age-specific infection fatality rates (IFR) and 95% confidence intervals. Infection fatality rates were obtained from [19].

| Age Group | % of Deaths | Mean IFR (%) | 95% CI |
| --- | --- | --- | --- |
| 0 – 19 | 0.0403 | 0.002 | 0.001 - 0.005 |
| 20 – 29 | 0.1500 | 0.0055 | 0.002 - 0.012 |
| 30 – 39 | 0.5113 | 0.024 | 0.007 – 0.046 |
| 40 – 49 | 1.6839 | 0.075 | 0.022 - 0.140 |
| 50 – 59 | 4.8405 | 0.207 | 0.062 - 0.388 |
| 60 – 69 | 8.9446 | 0.456 | 0.138 - 0.860 |
| 70 – 79 | 23.5888 | 1.674 | 0.502 – 3.139 |
| 80+ | 60.2604 | 8.292 | 2.488 - 15.547 |

**Table 2.**
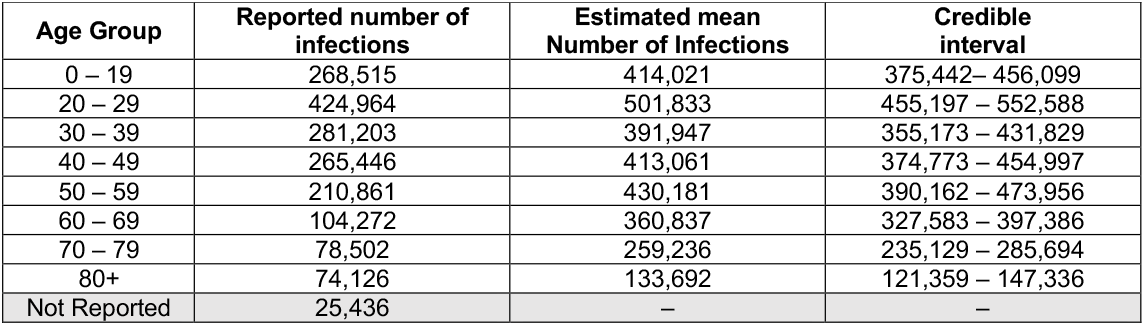
Reported cases and estimated number of SARS-CoV-2 infections in Japan by age groups.

| Age Group | Reported number of infections | Estimated mean Number of Infections | Credible interval |
| --- | --- | --- | --- |
| 0 – 19 | 268,515 | 414,021 | 375,442– 456,099 |
| 20 – 29 | 424,964 | 501,833 | 455,197 – 552,588 |
| 30 – 39 | 281,203 | 391,947 | 355,173 – 431,829 |
| 40 – 49 | 265,446 | 413,061 | 374,773 – 454,997 |
| 50 – 59 | 210,861 | 430,181 | 390,162 – 473,956 |
| 60 – 69 | 104,272 | 360,837 | 327,583 – 397,386 |
| 70 – 79 | 78,502 | 259,236 | 235,129 – 285,694 |
| 80+ | 74,126 | 133,692 | 121,359 – 147,336 |
| Not Reported | 25,436 | – | – |

To account for the uncertainty in the IFR estimates, we sampled IFR values from age-specific log-normal distributions, with the mean parameters informed from published studies [19]. The scale parameter of the log-normal distribution was set to 0.1, relatively large compared to the mean estimates, to allow for a larger range of the IFR estimates to be sampled.

Next, we estimated the likely date of infection for each inferred infection, regardless of whether the patient recovered or died. Specifically, for each infection, we estimated the lag-time *t*′ such that the infection occurred *t* − *t*′ days before the reported death on day *t*. This lag-time was calculated as the sum of two independent intervals: the incubation period and the time-to-death following symptom onset. These two intervals were sampled for each inferred infection to account for uncertainty; in particular, the incubation period was sampled from a log-normal distribution with mean 5.7 days and standard deviation of 3.1 days [20] and the time-to-death was sampled from a lognormal distribution with mean of 14.5 days and a standard deviation of 6.7 days [21]. The total number of infections on any given day *τ* is then calculated by summing all infections associated to deaths at time *t* > *τ* (i.e., *N*_*t*_) where the infection lag-time *t*′ is such that *t* − *t*′ = *τ*.

To validate our estimates, we conducted a meta-analysis of multiple serology reports from January 1, 2020, to December 31, 2020 [22–24]. Study eligibility criteria were population-based studies describing the prevalence of anti-SARS-CoV-2 (IgG and/or IgM) serum antibodies. Participants were from different socioeconomic backgrounds and age groups. A random-effects model was used to estimate pooled seroprevalence, and then extrapolated to the general population in June 2020 and December 2020. Heterogeneity was assessed using I^2^ statistics. Subgroup analyses were performed to explore potential sources of heterogeneity in the data.

The variation in parameters was accounted for by running 1000 Monte Carlo simulations. The results are summarized by calculating the mean and associated credible interval of the inferred number of infections. All relevant code and data files are available on https://github.com/BlueDot-global/true_infections.

## Results

We estimate that as of December 31, 2021, 2.90 million (CrI: 1.77 to 4.27 million) people have been infected in Japan, which is 1.68 times higher than the 1.73 million reported cases, resulting in a case ascertainment rate of 59.7% (CrI: 40.5 to 97.6%) (Figure 1A). The estimated 2.90 million infections correspond to approximately 2.29% of the population of Japan. Stratified by age, we found the highest under-ascertainment in adults aged 60 – 69. Among this age group, we estimated a total of 360.8 thousand infections (CrI: 327.6 – 397.3 thousand), of which only 104.3 thousand were reported. For adults aged 20 – 29 and 50 – 59, we estimated 501.8 thousand infections (CrI: 455.2 – 553.6 thousand) and 430.2 thousand infections (CrI: 390.2 to 455.0 thousand), respectively. Total number of infections among children between 0 – 19 years of age was estimated to be 414.0 thousand (CrI: 375.4 – 456.1 thousand). Figure 1B shows the number of estimated number of infections by age, overlaid with reported case counts.

**Figure 1.**
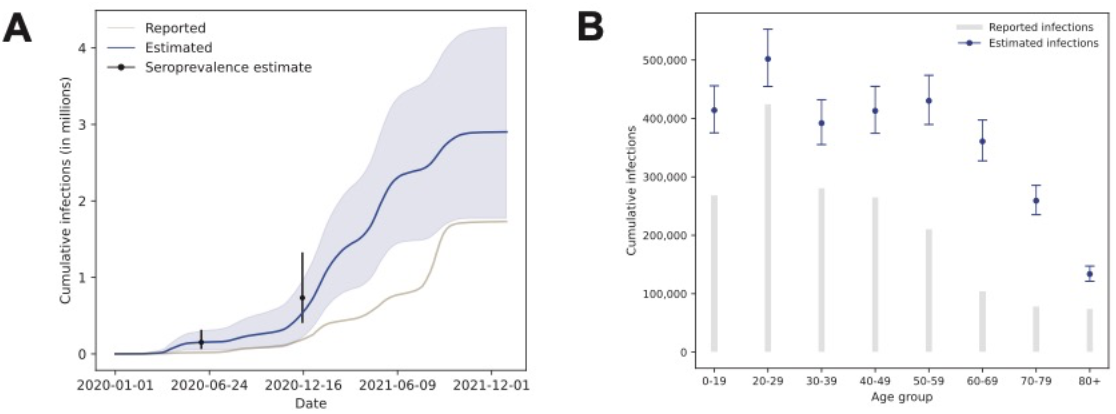
(A) Estimated total number of SARS-CoV-2 infections in Japan from January 16, 2020 to December 31, 2021. Black bars represent cumulative number of infections inferred by seroprevalence studies [22–24]. (B) Estimated total number of infections, stratified by age group.

Our estimates of the total number of infections are consistent with multiple serology reports. Using a random-effects model, we estimated the pooled seroprevalence of the general population in June 2020 and December 2020 to be 0.12% (95% confidence interval (CI): 0.05% - 0.25%) and 0.58% (95% CI: 0.32% - 1.05%), respectively. In comparison, the mean result of our estimation showed that 0.12% of the population was infected as of June 2020 and 0.43% as of December 2020, showing significant agreement with clinically observed serology estimates.

## Discussion

Understanding the extent of unreported infections in a country is crucial for effective policy and mitigation strategies and estimating the severity of the outbreak. In this study, we estimated the level of case ascertainment in Japan using a statistical framework that combines IFR estimates and publicly reported death counts. Our results indicate that a substantial number of COVID-19 infections in the country had not been reported, particularly in adults aged 60 to 69 years of age. This under-ascertainment is explained by two crucial factors. First, early in the pandemic, Japan employed a RT-PCR testing policy in which individuals with mild infection were only eligible for testing if their symptoms persisted for 4 days or longer [25]. Indeed, Japan had only tested 0.2% of its population by May 2020, the lowest rate among high-income, developed countries [25,26]. This policy was eventually amended in late 2021 to make antigen and PCR tests available for asymptomatically infected individuals [27]. Second, by the end of 2021, the vaccination rate had exceeded 80% [28]. Since vaccines prevent COVID-19 disease but not SARS-CoV-2 infection, increasing coverage shifts symptomatic infections to asymptomatic infections [29], thereby exacerbating the under-ascertainment.

Despite the relatively less robust testing strategies, social distancing measures, and lockdown restrictions, the estimated prevalence of 2.29% by end of 2021 was lower than other countries in the Western Pacific as well as USA and the UK [30]. Japan’s high population density and early exposure to SARS-CoV-2 make this pattern more counterintuitive. This low prevalence can be explained by several reasons. First, due to the already established social norm, the public’s compliance to wearing face masks and coverings during COVID-19 was strong both early and late stages in the pandemic [31]. In addition, considering shortages of medical masks in the early days of the pandemic, the government sent over 100 million reusable face masks to its citizens in April 2020 ensuring strong compliance early in the pandemic [32]. Second, Japan established a robust network of contact tracers to track the spread of SARS-CoV-2 [33]. Considering that roughly 20% of SARS-CoV-2 infections are responsible for 80% of transmission [34], the country adopted cluster-focused retrospective contact-tracing methodology; this method looks backwards to identify when and where a case was originally infected to isolate COVID-19 transmission clusters. Published studies have shown that retrospective contact tracing results in identifying 2 - 3 times more cases than prospective contact tracing [35]. Finally, we presume that efficacious messaging on the risk factors of airborne viral transmission early in the pandemic, along with the culture of shared responsibility, lead to strong self-regulation and awareness in the population.

Our estimates of case ascertainment should be considered in the context of several limitations. While death counts are a more accurate indicator of disease burden [15], the number of deaths attributable to COVID-19 may have been underestimated, especially during the first months of the pandemic, affecting our model estimates [16]. Further, IFR values are subject to considerable change due to factors that affect severe outcomes such as increasing vaccination coverage and emerging new variants in the population. Specifically, we note that all IFR values were obtained from seroprevalence studies conducted before vaccination. Nonetheless, our estimates are consistent with serological data from Japan, with model’s mean estimates considerably overlapping the 95% confidence intervals reported in the studies.

Effective vaccination strategies to achieve herd immunity and non-pharmaceutical control measures, coupled with robust surveillance systems, are needed to restrain the spread of SARS-2-CoV variants in the country. Sustainable and feasible long-term control of COVID-19 requires continuous review and understanding of the severity of the ongoing pandemic. Our method to estimate the true underlying cases could be a fundamental component to strengthen these population health control and surveillance measures.

## Data Availability

All data produced in the present study are available upon reasonable request to the authors and available online at https://github.com/BlueDot-global/true_infections

## Conflict of interest/disclosure

KK is the founder of BlueDot, a social enterprise that develops digital technologies for public health. LM, AS, AD, KW, and KK are employed at BlueDot. KI and EY are employed at Meiji Seika Pharma.

## Author Contributions

AS, LM, AD, and KK conceived the idea. LM, KI, and EY collected and contributed data. LM, AS, and KW performed statistical analysis and code programming. All authors interpreted the results and contributed to the writing of the manuscript.

## Notes

### Competing Interest Statement

Kamran Khan is the founder of BlueDot, a social enterprise that develops digital technologies for public health. Lauren McKenzie, Affan Shoukat, Alex Demarsh, Kai On Wong, and Kamran Khan are employed at BlueDot.
Koju Itahashi and Eiji Yasuda are employed by Meiji Seika Pharma Co.

### Funding Statement

This study did not receive any funding

